# Fewer Boys Were Born After Brazil’s Gamma COVID-19 Wave and Pelé’s Death: A National Time-Series Analysis

**DOI:** 10.64898/2026.09.28.26364140

**Authors:** Debora F.B. Leite, Rumbidzai Masukume, Pugie T. Chimberengwa, Victor Grech, Witness Mapanga, Gwinyai Masukume, Grażyna Liczbińska

## Abstract

The sex ratio at birth (SRB), or secondary sex ratio, is stable, with more boys born alive, but shifts after major events. Acute stress may reduce SRB through disproportionate male fetal loss, while sexual intercourse patterns around conception are associated with offspring sex. We examined whether Brazil’s April 2021 Gamma-wave mortality peak was followed by an SRB decline within the previously reported stress-sensitive 3–5-month window. We analysed >81 million live births (1997–2024) from Brazil’s Live Birth Information System (SINASC) using pre-pandemic autoregressive forecasts with 95% prediction intervals (PIs). In July 2021, SRB was 50.951% versus 51.185% expected, below the 95% PI (50.968–51.402%), indicating fewer boys. All five macroregions were below expectation, largest in the North, including Amazonas, where Gamma was first identified. Monte Carlo simulation showed that, under usual SRB patterns, a decline with this timing and consistency across Brazil would occur about once in 200 simulations (probability 0.00467), making July 2021 unusual. In February 2023, SRB was 51.441% versus 51.220% expected, above the 95% PI (51.003–51.438%), indicating more boys, nine months after Brazil ended its COVID-19 emergency, suggesting increased sexual intercourse after pandemic relaxation. In April 2023, SRB was 50.956% versus 51.221% expected, below the 95% PI (51.004–51.439%), indicating fewer boys, four months after Pelé’s death. A comparable UK decline followed Princess Diana’s death 4–5 months later. Gamma was prespecified; later observations are hypothesis-generating. These findings highlight SRB’s population-level utility and warrant investigation of stress-related and behavioural pathways; ecological design precludes causality.

## Introduction

The sex ratio at birth (SRB), also known as the secondary sex ratio, usually expressed as the proportion of male live births among all live births, is remarkably stable at population level, with a modest male excess. Nevertheless, short-lived departures have been reported after wars, natural disasters, economic shocks, terrorist attacks, epidemics, and other acute population stressors [1–3]. One proposed pathway is disproportionate loss of vulnerable male fetuses after acute stress, producing fewer male live births several months later. Male fetuses may be more susceptible to adverse intrauterine conditions because of sex-specific differences in developmental trajectories and placental or physiological responses to maternal stress. This proposed vulnerability may also reflect the greater energetic demands of carrying a male fetus; women carrying male fetuses have been reported to have higher energy intake, and male newborns are, on average, heavier at birth [4].

An early COVID-19 hypothesis similarly proposed that pandemic-related stress could lead to a decline in SRB through greater male fetal vulnerability [5]. This raises a broader epidemiological question: whether SRB can show a measurable, time-lagged response to major population-level stressors. This literature is often discussed in relation to the Trivers–Willard hypothesis, although the applicability and mechanism of Trivers–Willard effects in humans remain debated [6–8]. Importantly, an observed population-level change in SRB does not itself establish the Trivers–Willard mechanism, which remains one possible biological explanation among several.

The World Health Organization characterized COVID-19 as a pandemic on 11 March 2020 [9], providing an opportunity to examine the relationship between a major population-level health shock and a subsequent reproductive outcome. Significant transient SRB declines were reported three months after pandemic declaration in South Africa and in England and Wales [10, 11]. Northern Ireland showed a decline five months after the pandemic declaration, while the Republic of Ireland and Japan showed declines nine months later [12–14]. A later Japanese national analysis also identified anomalous male-birth proportions during 2020–2022, including the previously reported December 2020 decline [15]. South Korea showed a nationwide decline during the pandemic, with a pronounced trough in Daegu approximately nine months after the initial outbreak peak [16]. In contrast, nationwide analyses from the United States, Croatia, and Finland found no overall COVID-19-related SRB perturbation [3, 17, 18]. An analysis from Western Sydney, Australia, similarly found no overall SRB change after pandemic onset, although subgroup-specific variation was observed by maternal country of birth [19]. The evidence is therefore heterogeneous rather than uniformly positive or null, with reported findings differing in magnitude and, importantly, in timing.

The United States study is particularly relevant because Cleaver and Non [17] explicitly tested the Trivers–Willard hypothesis in more than 25 million singleton births and found no national change in SRB. This finding is an important counterpoint to studies reporting transient declines and shows that a large population denominator alone does not guarantee detection of an SRB response. The contrast also highlights the potential importance of population characteristics, epidemic trajectory, and mortality burden when assessing whether a national SRB signal is detectable.

Brazil provides an unusually informative setting. It has a population exceeding 200 million people, pronounced regional and socioeconomic heterogeneity, and a consistent national live-birth information system with millions of births annually. These characteristics provide both a large denominator for detecting relatively small changes in SRB and an opportunity to assess whether a national signal is geographically coherent. The country experienced one of the world’s most severe COVID-19 mortality burdens and, crucially for the present hypothesis, a major early-2021 wave associated with the Gamma (P.1) lineage, which was first identified in Manaus, Amazonas and later spread nationally [20, 21]. Reported national COVID-19 related deaths peaked in April 2021 [22]. A transient SRB decline approximately 3–5 months later would therefore be temporally consistent with the lag observed in several previous population-stress and COVID-19 studies [10, 11, 14].

Brazil also offers two valuable contextual checks. First, a prior analysis of approximately 65.8 million births from 1997–2018 established a strong March–May birth peak, corresponding broadly to conceptions during the Southern Hemisphere winter [23]. Second, Brazil experienced the 2015–2016 Zika epidemic. Independent demographic studies documented a reduction in fertility and live births after recognition of the association between Zika virus infection and congenital abnormalities [24–27]. Thus, a Zika-era trough in the same series can function as an internal contextual validation feature: not as evidence for a particular COVID-19 mechanism, but as an indication that the national birth series can capture a previously documented, temporally localized reproductive response to a major population health event.

Against this background, Brazil provides an opportunity to examine whether a clearly defined population-level mortality shock was followed by a transient SRB deviation within a prespecified temporal window, and whether any such deviation showed geographic coherence. The aim of this study was to determine whether Brazil’s severe COVID-19 mortality wave was followed by an unusual decline in national SRB 3–5 months after the April 2021 Gamma-wave mortality peak. In addition, we examined whether the direction and timing of the national SRB deviation were consistent across Brazil’s five macroregions. We also examined the broader temporal pattern of SRB during 2020–2024 to identify deviations occurring outside the prespecified window.

## Methods

### Data source

Monthly recorded live births were obtained from Brazil’s *Sistema de Informações sobre Nascidos Vivos* (SINASC), Ministry of Health. The available file extended back to 1994, but the primary study period began in January 1997 because national completeness and comparability of SINASC data are considered more suitable from this period onward, and because the principal published analysis of Brazilian birth seasonality used the same starting year [23, 28–30]. The dataset contained monthly counts of male and female live births for Brazil as a whole and for its five macroregions. The study used aggregate, publicly available data and no individual-level or identifiable information. Ethics committee approval and informed consent were therefore not required. All statistical analyses were performed using Stata/BE version 17.0 (StataCorp LLC, College Station, TX, USA).

### Birth seasonality and Zika context

Monthly birth counts were converted to mean births per day to account for unequal month length. Within each calendar year, months were ranked from 1 (highest mean daily births) to 12 (lowest). Approximate conception-month ranks were displayed by shifting birth-month ranks nine months earlier; this is an ecological approximation and does not assign individual conception dates.

The seasonality analysis was used as a contextual check: we assessed whether the previously reported March–May Brazilian birth peak persisted through 2024. Rank distributions for March, April, and May were compared between 1997–2019 and 2020–2024 using Wilcoxon rank-sum tests. We treated the Zika-era birth trough as an internal contextual feature of the longitudinal series. This was not a second causal analysis, and no Zika-specific time-series model was fitted. Rather, we assessed whether the series showed the previously documented, temporally localized decline in recorded births following the emergence of Zika-associated congenital abnormalities [24–27]. This analysis was intended as a contextual check on the sensitivity of the national birth series to a major population health event, rather than as a test of a common causal mechanism.

### Sex ratio at birth

The SRB was calculated each month as male live births divided by total live births and expressed as a percentage. The primary analysis used a parsimonious autoregressive moving-average model with one autoregressive term [ARMA(1,0)]. The model was fitted to the recent pre-pandemic baseline from January 2015 through December 2019 (60 monthly observations). A five-year baseline was chosen in line with previous SRB studies and COVID-19 forecasting practice [13, 14, 16]. Stationarity of the baseline SRB series was assessed with the augmented Dickey–Fuller test.

Model coefficients estimated from the pre-pandemic period were held fixed. Beginning in January 2020, a rolling one-step-ahead forecast was generated for each month: after the forecast was made, the observed SRB for that month was appended to the series and used to condition the forecast for the following month, without re-estimating the ARMA coefficients. Thus, each forecast used only information that would have been available immediately before the month being forecast, while the underlying model parameters remained fixed at their pre-pandemic estimates. Ninety-five percent prediction intervals were obtained from the forecast distribution. The biological hypothesis concerned a short-lived reduction in SRB after severe population stress.

Monthly reported confirmed COVID-19 deaths for Brazil were obtained from Our World in Data [22] and aggregated by calendar month for temporal comparison with SRB. The national mortality maximum occurred in April 2021 during the Gamma-dominant wave. We therefore examined whether SRB fell below its expected range approximately 3–5 months later, corresponding to the hypothesized lag between a major population stressor and a subsequent change in the sex composition of live births. An observed SRB below the lower 95% prediction bound was considered unusual relative to the pre-pandemic process. The direction and timing of any departure were interpreted together; an isolated crossing of a prediction limit was not interpreted as evidence of a causal effect, particularly given the number of monthly forecasts examined. As a post hoc exploratory analysis, the same modelling approach was applied separately to Brazil’s five macroregions (North, Northeast, Southeast, South, and Central-West). We examined whether the July 2021 deviation was concordantly downward across regions, and whether the largest deviation occurred in the North, which includes Amazonas, the state in which the Gamma (P.1) lineage was first identified [20, 21]. These regional analyses were used to assess geographic coherence of the national signal and were not treated as independent confirmatory tests. Monte Carlo simulation was used to estimate the probability of observing the combined temporal and geographic pattern under the pre-pandemic error structure. Specifically, the joint distribution of the national and five macroregional one-step-ahead forecast errors was estimated from the 60 monthly observations in 2015–2019. The covariance matrix was calculated from these contemporaneous forecast errors, and correlated errors were generated from a six-dimensional multivariate normal distribution with zero means and the estimated covariance matrix. A total of 2,000,000 simulations were generated using a fixed random seed (20260909). Pre-pandemic forecast errors were also assessed for first-order serial correlation using the Ljung–Box test. For each simulation, the same temporal and geographic criteria used for the observed data were applied. The complete configuration for a single month was defined as a national forecast error at least as negative as that observed in July 2021, negative forecast errors in all five macroregions, and the North having the most negative regional deviation. The empirical single-month probability was calculated as the proportion of simulations meeting all three criteria. The probability of observing the complete configuration at least once within the three possible months corresponding to the prespecified 3–5-month interval after the April 2021 mortality peak (July–September 2021) was then calculated as 1 − (1 − p)^3. This calculation assumes independence of the monthly one-step-ahead innovations; the absence of first-order serial correlation was assessed using the Ljung–Box diagnostic. The resulting probability was interpreted as an exploratory measure of how unusual the observed joint temporal and geographic configuration was under the specified pre-pandemic error structure, rather than as a conventional confirmatory p-value. For the broader 2020–2024 series, months outside the 95% prediction interval but outside the prespecified Gamma window were retained as exploratory observations. Their temporal relation to later population events was considered only after the deviations had been identified and was therefore treated as retrospective, hypothesis-generating context rather than confirmatory testing. No additional event-specific probability was calculated for these later observations.

### Annual recorded live births

Annual totals were obtained by summing monthly births. For descriptive context, a linear regression of annual births on calendar year was fitted over 1997–2019 and projected through 2024 with 95% confidence intervals. This model was intended as a transparent historical benchmark rather than a structural fertility forecast. Annual recorded births were not interpreted as fertility rates because they also depend on population size and age structure.

### AI use disclosure

ChatGPT (OpenAI) was used for language editing and to assist the authors with statistical code development and verification. The authors independently reviewed the analyses and outputs against the source data and take full responsibility for the final manuscript.

## Results

### Birth seasonality and Zika context

Across 1997–2024, the established Brazilian seasonal pattern was highly persistent. March was the highest-ranked birth month in 15 years, April in nine years, and May in four years; no other month ranked first (Fig. 1). The corresponding approximate conception pattern was concentrated in June–August (Fig. 1). There was no evidence of a reorganization of this seasonal pattern during the COVID-19 period: rank distributions for March (p = 0.529), April (p = 0.799), and May (p = 0.616) did not differ between 1997–2019 and 2020–2024.

**Fig. 1.**
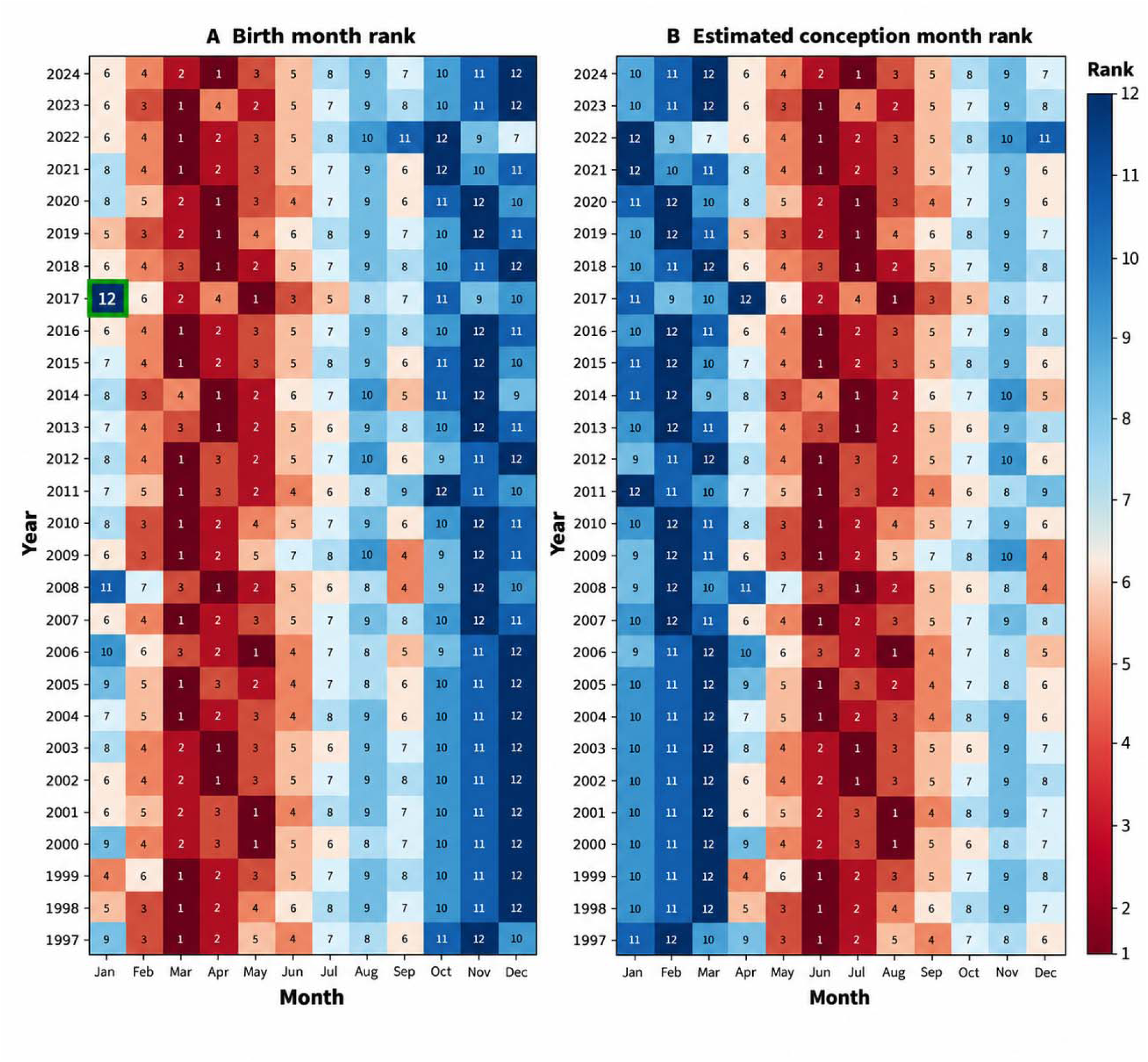
Seasonal birth and approximate conception patterns in Brazil, 1997–2024. (A) Birth months ranked by mean daily live births within each year (1 = highest; 12 = lowest); January 2017 is outlined as the internal contextual feature. (B) Approximate conception-month ranks, obtained by shifting birth-month ranks nine months earlier.

January 2017 was a conspicuous exception to the usual trough pattern. It averaged 7,302 recorded live births per day and was the lowest-ranked month of 2017; it was the only January to rank 12th during the entire 28-year study period. The timing of this trough is consistent with previously documented reductions in fertility and live births following the 2015–2016 Zika epidemic.

### Sex ratio at birth

The January 2015–December 2019 monthly SRB series was stationary (augmented Dickey– Fuller statistic = −5.941, p < 0.001). The fitted ARMA(1,0) model estimated a mean SRB of 51.214% and an autoregressive coefficient of 0.247. Residual diagnostics did not indicate remaining first-order autocorrelation. During 2021, the national monthly SRB remained within the rolling 95% prediction interval except in July (Table 1; Fig. 2). The observed July 2021 SRB was 50.951%, compared with an expected value of 51.185% (95% prediction interval, 50.968%– 51.402%). Thus, the observed SRB was 0.234 percentage points below the expected value and below the lower prediction limit.

**Fig. 2.**
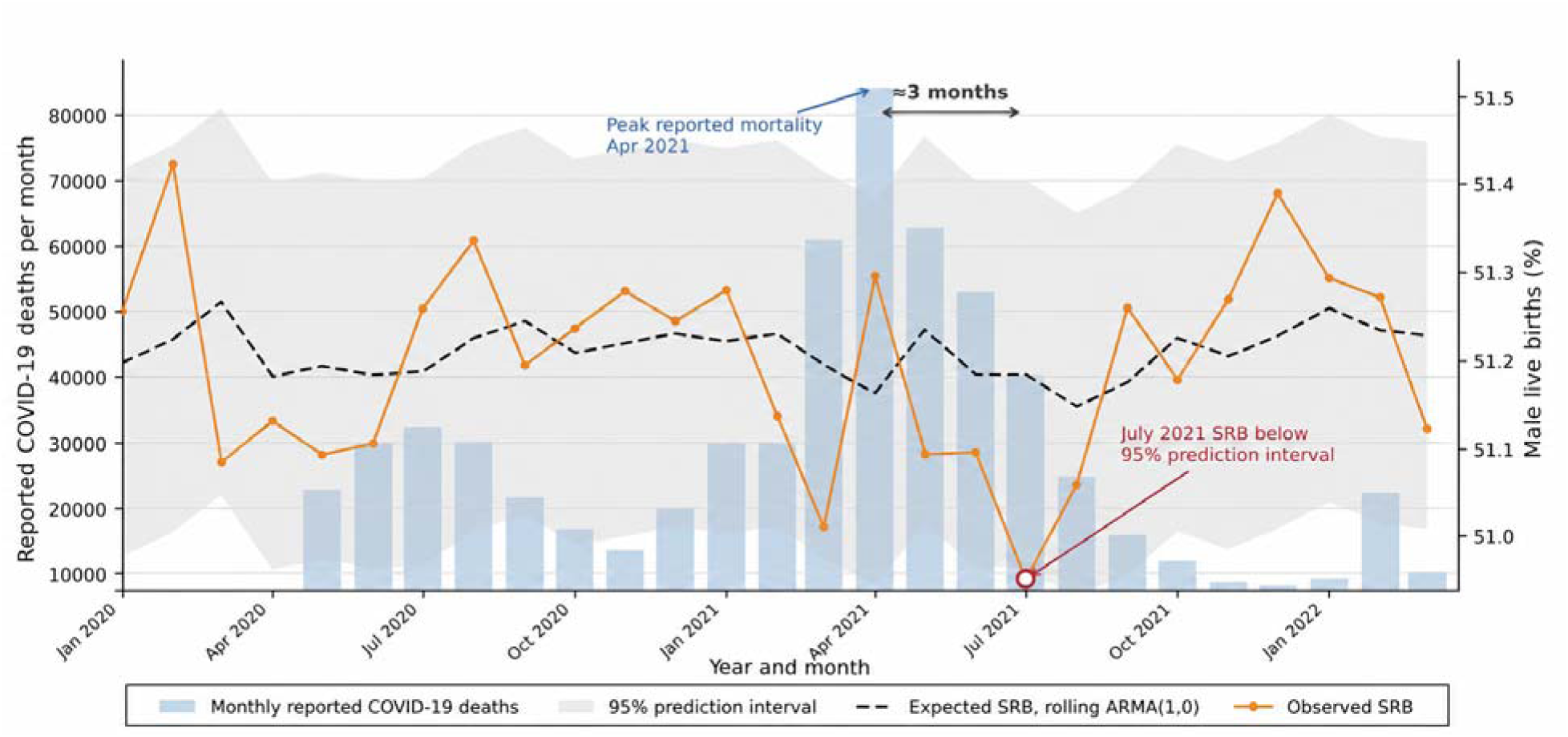
COVID-19-related deaths and sex ratio at birth in Brazil, January 2020 to March 2022. Monthly reported COVID-19 deaths are shown alongside observed and predicted SRB with 95% prediction intervals. The July 2021 SRB fell below the lower prediction limit three months after the April 2021 mortality peak.

**Table 1.** Observed and predicted national sex ratio at birth, Brazil, 2021.

| Month | Male | Female | Observed SRB (%) | Predicted SRB (%) | 95% PI (%) | Outside 95% PI |
| --- | --- | --- | --- | --- | --- | --- |
| January | 113,831 | 108,152 | 51.279 | 51.222 | 51.004–51.439 | No |
| February | 108,294 | 103,482 | 51.136 | 51.230 | 51.013–51.448 | No |
| March | 122,933 | 118,062 | 51.011 | 51.195 | 50.978–51.412 | No |
| April | 119,562 | 113,523 | 51.295 | 51.164 | 50.947–51.381 | No |
| May | 122,577 | 117,331 | 51.093 | 51.234 | 51.017–51.452 | No |
| June | 113,534 | 108,663 | 51.096 | 51.184 | 50.967–51.402 | No |
| July | 113,475 | 109,238 | 50.951 | 51.185 | 50.968–51.402 | Yes |
| August | 112,107 | 107,456 | 51.059 | 51.149 | 50.932–51.367 | No |
| September | 113,234 | 107,670 | 51.259 | 51.176 | 50.959–51.393 | No |
| October | 110,140 | 105,070 | 51.178 | 51.225 | 51.008–51.443 | No |
| November | 108,239 | 102,884 | 51.268 | 51.205 | 50.988–51.423 | No |
| December | 111,632 | 105,595 | 51.390 | 51.228 | 51.010–51.445 | No |

Note. PI = prediction interval; SRB = sex ratio at birth. Prediction intervals are from rolling one-step-ahead forecasts using the fixed pre-pandemic ARMA(1,0) model.

Reported COVID-19 mortality rose sharply during the Gamma-dominant wave and reached its national monthly peak in April 2021 (Fig. 2). The July 2021 SRB deviation occurred three months after this mortality peak, within the hypothesized 3–5-month interval. The departure was transient, with the SRB remaining within the prediction interval in the adjacent months.

The complete January 2020 to December 2024 observed and forecast SRB series is shown in Fig. 3; the additional observations outside the 95% prediction interval are described below.

**Fig. 3.**
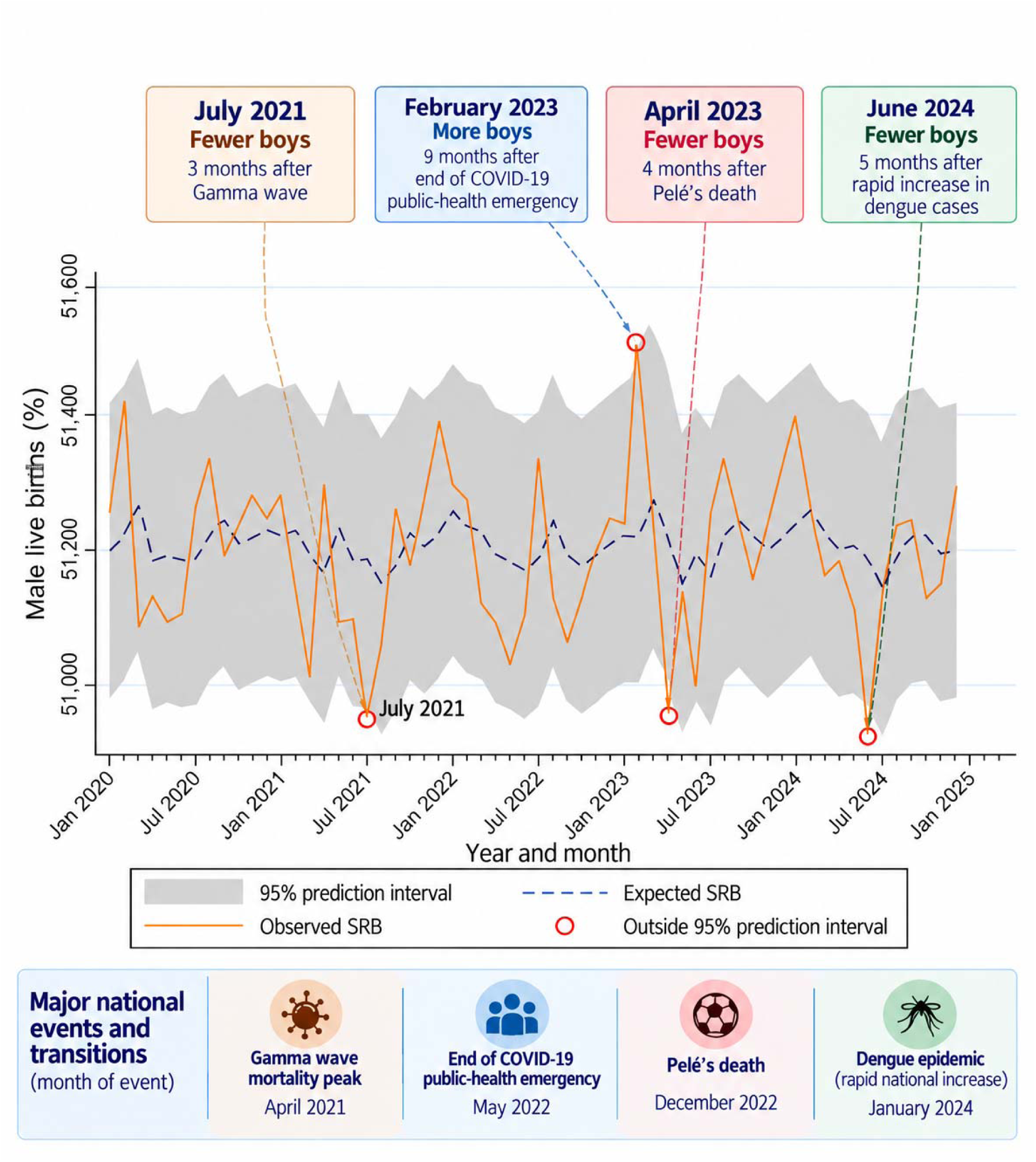
National sex ratio at birth in Brazil, 2020–2024. Observed and expected monthly SRB with 95% prediction intervals. Circles denote observations outside the interval; annotations identify the July 2021, February and April 2023, and June 2024 observations discussed in the text.

As a post hoc exploratory macroregional analysis, the July 2021 SRB was below expectation in all five macroregions (Fig. 4). The largest deviation occurred in the North (−0.414 percentage points), while the Southeast contributed the largest absolute number of births to the national deviation because of its larger birth volume. No individual macroregion crossed its 95% prediction interval. The probability of observing the complete specified configuration at least once within the three possible months corresponding to the prespecified 3–5-month interval after the April 2021 mortality peak (July–September 2021) was 0.00467 (about 1 in 200). The configuration required a national decline at least as large as that observed in July 2021, downward deviations in all five macroregions, and the largest regional deviation in the North. This was treated as an exploratory measure of the unusualness of the joint pattern under the pre-pandemic error structure, rather than as a conventional confirmatory p-value.

**Fig. 4.**
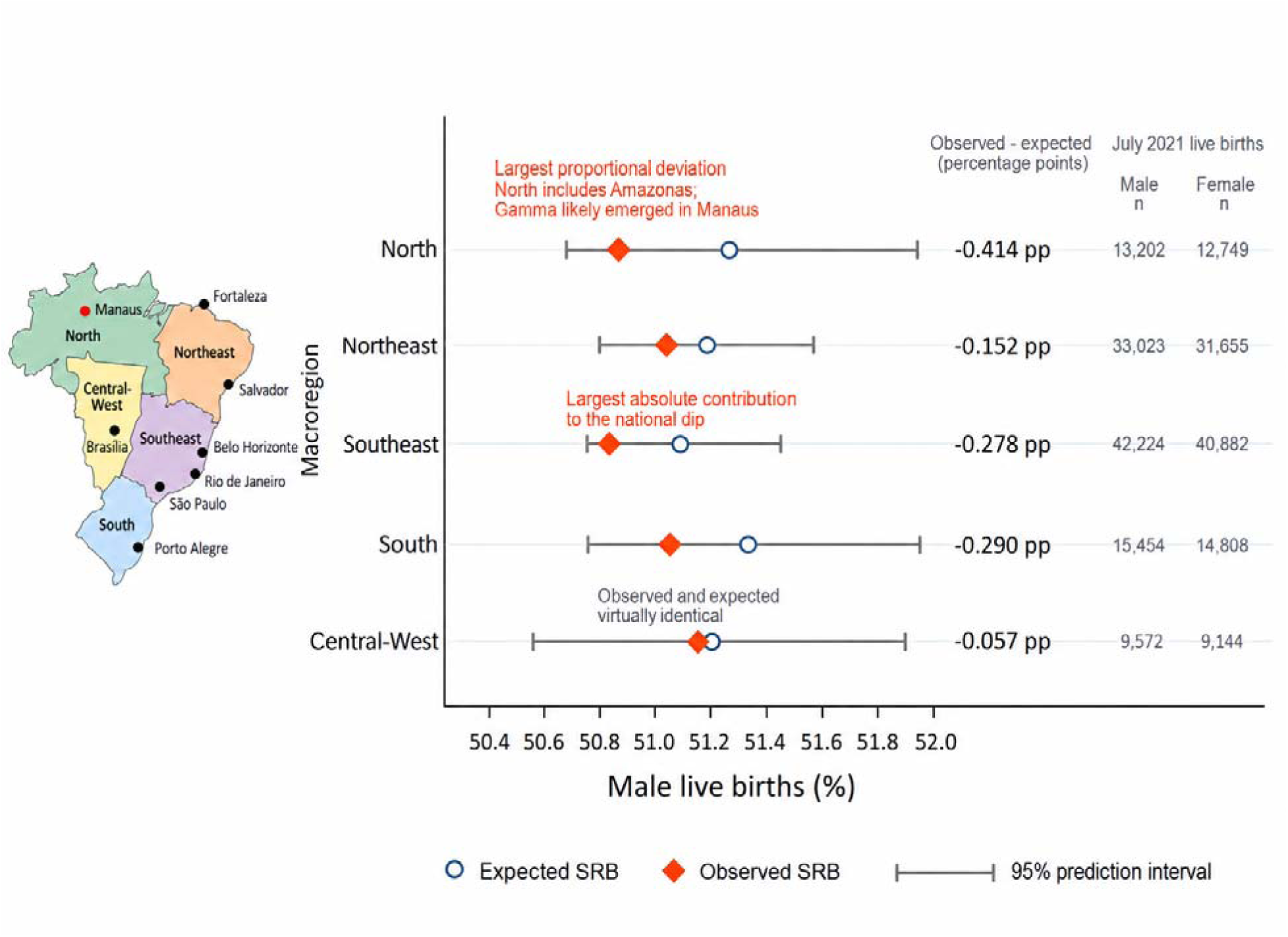
Sex ratio at birth by Brazilian macroregion in July 2021. Observed and expected percentages of male live births and region-specific 95% prediction intervals are shown for each macroregion.

Three additional national observations fell outside the rolling 95% prediction interval during 2020–2024 (Fig. 3). In February 2023, SRB was 51.441% versus 51.220% expected (95% PI, 51.003%–51.438%), a 0.221 percentage-point upward deviation, nine months after Brazil ended its COVID-19 public-health emergency in May 2022. In April 2023, SRB was 50.956% versus 51.221% expected (95% PI, 51.004%–51.439%), a 0.265 percentage-point downward deviation, approximately four months after Pelé’s death on 29 December 2022. In June 2024, SRB was 50.921% versus 51.188% expected (95% PI, 50.971%–51.406%), a 0.267 percentage-point downward deviation. These observations were outside the prespecified Gamma-wave hypothesis and are exploratory.

### Annual recorded live births

Annual recorded live births were approximately 3 million in the late 1990s and early 2000s, reaching 3,250,433 in 1999, followed by a long-term decline with intermittent rebounds (Fig. 5). A linear regression fitted to annual births over 1997–2019 predicted 2,839,232 births in 2020 (95% CI, 2,772,071–2,906,392). The observed number was 2,729,701 and observed annual totals remained below the lower 95% confidence limit of the historical benchmark through 2024. In 2024, 2,388,959 births were recorded, compared with 2,848,712 in 2019, representing a 16.1% decline.

**Fig. 5.**
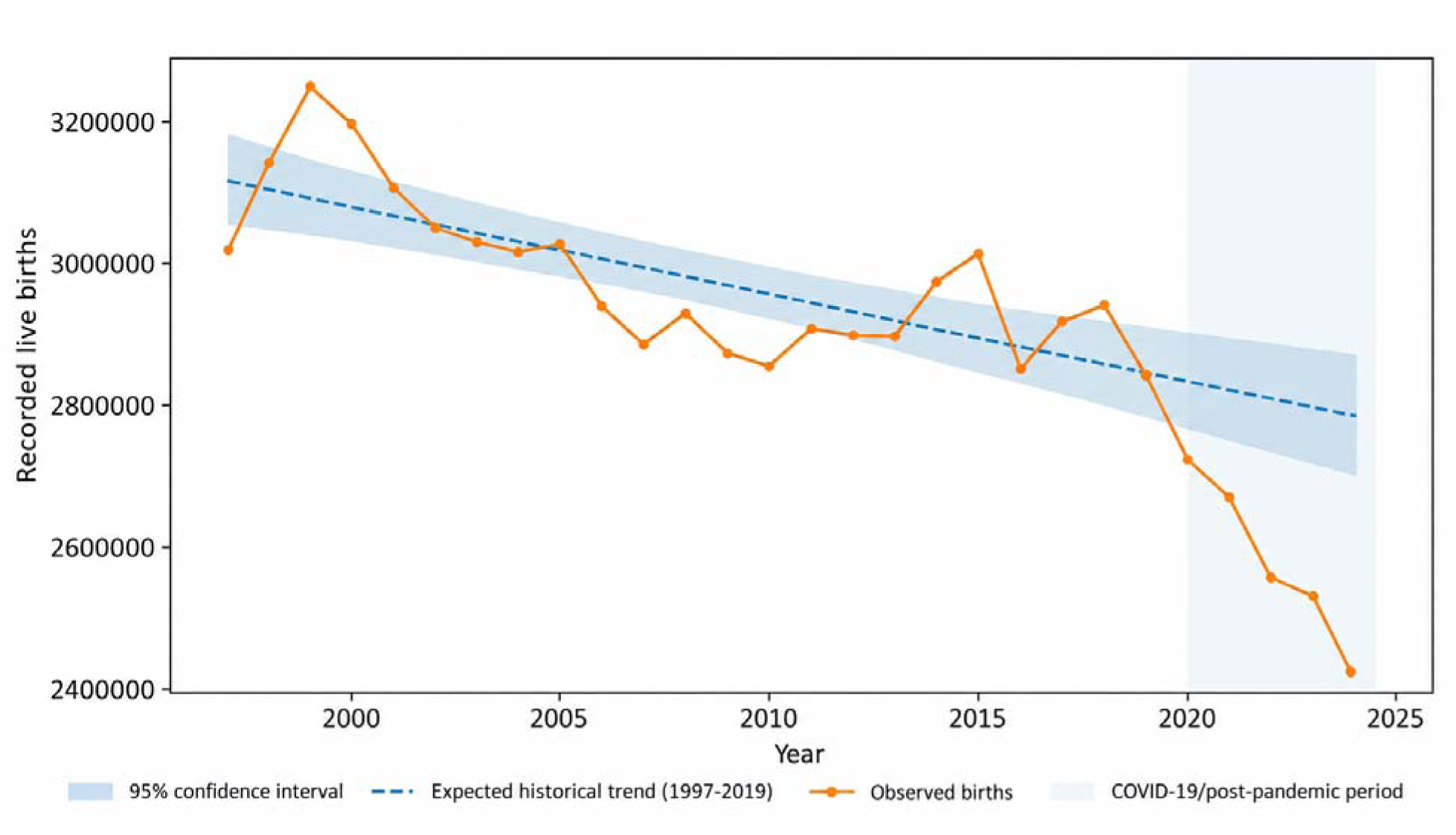
Annual recorded live births in Brazil, 1997–2024. Observed annual totals are shown with the 1997–2019 linear historical benchmark and its 95% confidence intervals.

## Discussion

Our results have confirmed our hypothesis: the principal finding is a national decline in Brazil’s SRB in July 2021. The observed SRB of 50.951% fell below the lower 95% prediction bound, approximately three months after the April 2021 mortality peak during the Gamma-dominant wave. This temporal relationship is consistent with the short-term population-stress pattern reported in several previous studies of sex ratio at birth, although it does not identify the biological or behavioural pathway responsible for the Brazilian observation. July 2021 was the only month in 2021 in which the observed national SRB fell outside the 95% prediction interval, which is consistent with a transient rather than sustained departure from the underlying SRB process.

The Brazilian result fits within a substantial but heterogeneous international literature. South Africa showed a significant June 2020 decline and inversion three months after pandemic onset; England and Wales showed a significant June 2020 decline at three months and a later December increase; Northern Ireland showed a significant August 2020 decline at five months and a December increase; the Republic of Ireland showed a marked December 2020 decline; Japan showed a significant December 2020 decline nine months after the pandemic declaration; and South Korea showed a nationwide pandemic-period decline, with a pronounced November 2020 trough in Daegu approximately nine months after its initial outbreak peak [10–14, 16]. In other words, these studies suggest that when SRB perturbations have been observed during the COVID-19 period, their timing has not been uniform, ranging from approximately three to five and at nine months after a major pandemic-related reference point. Inoue and Mizoue also found that the Japanese decline was concentrated in prefectures more severely affected by COVID-19 [12], providing precedent for considering geographical heterogeneity alongside national time-series patterns. In Iran, SRB did not differ significantly from the pre-epidemic level during the first nine months after the beginning of the epidemic but was significantly higher during months 10–13 [31].

Evidence from later pandemic waves is also relevant. A recent South African analysis identified a May–July 2021 SRB depression within the 3–5-month window after the January 2021 Beta-wave mortality peak [32]. In Brazil, SRB exceeded the upper 95% prediction bound in February 2023, nine months after the national COVID-19 public-health emergency ended in May 2022 [33]; a later SRB elevation was also reported in South Africa [32]. Increased sexual intercourse after prolonged restrictions is one possible behavioural pathway. Intercourse timing and coital rate around conception have been associated with offspring sex and population SRB [34, 35]. The Brazilian timing is therefore compatible with, but cannot establish, a behavioural mechanism linking pandemic relaxation, intercourse patterns, and higher SRB. Because this observation was identified from the broader series, it is hypothesis-generating.

The April 2023 decline provides a second exploratory observation relevant to population stress. Pelé died on 29 December 2022, approximately four months before SRB fell below its lower 95% prediction bound. Peer-reviewed scholarship has examined Pelé as a figure in the construction of Brazilian national identity [36], supporting the plausibility of his death as a nationally salient event.

Collective bereavement also has a direct precedent in the SRB literature. In 29,293,240 UK live births, Grech reported a significant decline in the male proportion 4–5 months after the death of Diana, Princess of Wales (p = 0.046) [37]. The similar interval after Pelé’s death is compatible with a stress-related male fetal-loss hypothesis, but the association was identified retrospectively and cannot distinguish an event effect from chance or other contemporaneous exposures. The June 2024 decline has no comparably discrete explanation. Brazil’s record dengue surge was already evident in early 2024 [38, 39] and is a candidate for future study, particularly because maternal dengue has been associated with stillbirth [40]; however, evidence of sex-specific fetal survival is lacking, so dengue should not be interpreted as an explanation of the June deviation.

Not every country has shown this pattern. Cleaver and Non [17] found no overall COVID-19-related change in SRB among more than 25 million singleton births in the United States. Pavić [18] reported no predicted 3–5-or 9-month departure in Croatia despite concurrent pandemic and earthquake stressors. Helle *et al*. [3], using nationwide Finnish data through 2024, likewise found no evidence of a COVID-19-related change in the sex ratio of live births or stillbirths. A Western Sydney study also found no aggregate shift, although subgroup-specific variation by maternal country of birth was observed [19]. These findings are important because they argue against a universal COVID-19-related effect on SRB. They instead suggest that the magnitude, timing, or detectability of any perturbation may depend on the intensity and type of stressor, population composition, social context, or the timing of exposure relative to conception and gestation. Brazil is informative in this context because the July 2021 departure followed an unusually severe and temporally well-defined mortality wave rather than simply the onset of the pandemic.

Brazil is also especially informative because the relevant stressor was not simply “the pandemic” as a single March 2020 event. The P.1 (Gamma) lineage was identified in Manaus, Amazonas, during a major resurgence of SARS-CoV-2 transmission [20, 21], which was associated with substantial mortality and health-system pressure, followed by a national mortality peak in April 2021 [20, 21, 41]. The July 2021 SRB departure therefore occurred with a distinct later mortality shock that can be located in time independently of the birth data.

The geographic pattern provides additional context. All five Brazilian macroregions showed a downward deviation in July 2021, with the largest deviation in the North. The North includes Amazonas, where the P.1 lineage was first identified [20, 21]. The Monte Carlo analysis indicated that the probability of observing the complete temporal and geographic configuration at least once within the three possible months corresponding to the prespecified 3–5-month interval after the April 2021 mortality peak (July–September 2021) was 0.00467 under the specified pre-pandemic error structure. The configuration required a national decline at least as large as that observed in July 2021, downward deviations in all five macroregions, and the largest regional deviation in the North. The geographic concordance therefore supports the coherence of the observed pattern but should not be interpreted as an independent confirmatory test or as evidence of causality.

Independent Brazilian evidence also points to substantial psychosocial and reproductive strain during the pandemic. In a large Brazilian survey of 45,161 adults, 40.4% reported frequently feeling sad or depressed and 52.6% frequently feeling anxious or nervous [42]. National routine-data analyses subsequently estimated stillbirths to be 6.2% above expected in 2021 and maternal deaths 103.3% above expected, with particularly large maternal-mortality increases in March and June 2021 [43]. These findings are relevant because they place the April 2021 mortality peak within a broader period of substantial disruption to population well-being, pregnancy outcomes, and maternal health. They do not establish the pathway underlying the July SRB decline, and they should not be interpreted as evidence that psychosocial stress, maternal infection, or maternal mortality caused the observed change. Rather, they provide independent epidemiological context for the magnitude of disruption experienced during the period in which the SRB departure occurred.

Mechanistically, the most commonly invoked explanation for a 3–5-month decline is a change in fetal survival, potentially involving disproportionate loss of male fetuses following acute maternal or population stress. Male fetuses are, on average, more vulnerable to adverse intrauterine conditions, and selection in utero has been proposed to explain SRB shifts after sudden shocks [44]. Evidence from COVID-19-era studies of twin births provides additional, although indirect, support for the possibility of altered fetal survival during pandemic stress. In Norway, the monthly odds of a live-born twin among male births showed a 27% downward level shift beginning in May 2020 after pandemic onset [45]. In the United States, conception-cohort analysis subsequently found fewer male twin births than expected among cohorts exposed in utero early in the pandemic [46]. These studies do not establish that differential male fetal loss produced the Brazilian SRB decline, but they indicate that changes in fetal survival can accompany major population-level stress.

The observed direction is compatible with, but does not specifically support, a Trivers– Willard interpretation; the distinction between a population-level prediction and the specific evolutionary mechanism proposed by Trivers and Willard [8] is important. The original hypothesis proposes adaptive parental adjustment of offspring sex in relation to parental condition [8]. However, evidence for adaptive sex allocation in humans remains contested. Large-scale analyses have failed to provide consistent support for the hypothesis [6], while reviews of several decades of research conclude that the empirical evidence is heterogeneous and that proposed mechanisms remain difficult to distinguish [7]. The present study therefore does not provide evidence of adaptive parental sex allocation. Instead, it tests whether a population-level SRB perturbation occurred in the direction and temporal window expected under a broad population-stress hypothesis. Alternative pathways – including fetal loss unrelated to adaptive allocation, changes in the composition of pregnancies, maternal infection, changes in reproductive behaviour, or other unmeasured social and environmental exposures – remain possible [6, 7]. Our result consequently supports the observation of a transient population-level SRB response, not a specific individual-level evolutionary mechanism.

Two features of the longer Brazilian series provide useful contextual evidence regarding temporal structure and sensitivity to reproductive disruption. First, the March–May birth peak previously reported in Brazil persisted through 2024, showing that the data reproduce the established national seasonal pattern [23]. This is relevant because a stable seasonal structure reduces concern that the July 2021 observation simply reflects an unrecognised disruption of the broader annual birth cycle. Second, January 2017 was the only January in the 28-year series to rank as the annual birth trough. That timing is consistent with the independently documented reduction in fertility and live births during the Zika epidemic and following widespread recognition of Zika-associated congenital abnormalities [24–27]. The visibility of this reproductive perturbation in the same national series provides an internal contextual feature: the data are capable of capturing a temporally localized change in reproductive behaviour and birth numbers. It does not validate the ARMA counterfactual or prove a COVID-19 mechanism, but it provides supporting evidence that the longitudinal series retains sensitivity to major reproductive disruptions.

The continued decline in annual birth numbers provides broader demographic context but is not the core outcome. Brazil’s fertility transition long predates COVID-19, and Brazilian research has shown that reproductive responses to infectious-disease outbreaks can be heterogeneous across place and socioeconomic context [25, 26]. Zika itself altered reproductive behaviour and fertility in affected Brazilian populations [24, 25, 27], while subsequent work comparing Zika and COVID-19 documented different and potentially overlapping reproductive responses to successive infectious-disease outbreaks [26]. The decline from 2.85 million recorded births in 2019 to 2.39 million in 2024 should therefore be understood as part of a longer and heterogeneous fertility transition, rather than as a direct consequence of the pandemic.

## Strengths of the study

The study has several strengths. It uses national data from a population exceeding 200 million and more than 81 million recorded births over 1997–2024. The large number of births provides substantial statistical precision, while the 28-year series provide long temporal coverage and the national scope reduces the possibility that the principal finding is driven by a single locality. The SRB analysis uses a recent pre-pandemic baseline and rolling one-step-ahead forecasts, while the mortality series provides an independently timed population stressor rather than treating March 2020 as the only relevant pandemic exposure. The use of an independently derived mortality series also reduces reliance on the birth series itself to define the timing of the hypothesised stressor. This distinction is particularly important given evidence that SRB responses, when observed, may occur after different pandemic phases and at different lags [10–15]. The longer birth series also reproduces two established Brazilian features – the March–May seasonal peak and the Zika-era birth decline—and extends observation through 2024.

## Research limitations

There are several limitations to this study that affect the interpretation and generalizability of the findings. The analysis is ecological and includes all recorded live births rather than singleton births only. Individual maternal infection, psychosocial stress, socioeconomic position, parity, gestational age, “race”/“colour”, age, education, fetal loss, and vaccination status cannot be linked to offspring sex in the aggregate series. Consequently, the analysis cannot determine whether the national signal arose from changes in fetal survival, the composition of pregnancies, reproductive behaviour, maternal characteristics, or other correlated exposures. The national mortality peak also represents an ecological exposure measure and cannot establish that individual pregnant women experienced the same intensity or timing of COVID-19-related stress. Other contemporaneous changes, including reproductive behaviour, healthcare access, vaccination, infection prevalence, and fertility-related decisions, may have contributed to the observed temporal pattern and cannot be disentangled in the aggregate data. Because multiple monthly forecasts were examined, isolated crossings of the 95% prediction interval may occur by chance. This is particularly important for the February and April 2023 and June 2024 observations, which were identified from the broader series rather than specified in advance. The proposed links to pandemic relaxation and Pelé’s death are retrospective temporal correspondences and are vulnerable to event-selection and multiple-comparison bias; they should therefore be interpreted as hypothesis-generating rather than confirmatory. The July 2021 departure should likewise not be interpreted from prediction-interval crossing alone. Its evidential value rests on the combination of the magnitude and direction of the deviation, its temporal proximity to an independently observed mortality peak, its transient character, and the concordant direction across all five macroregions. The model was intentionally parsimonious.

The macroregional analysis and joint simulation were post hoc and exploratory; their value lies in assessing the geographical coherence of the national finding rather than providing an independent causal test. The Monte Carlo probability should therefore be interpreted as an exploratory measure of the unusualness of the observed joint pattern under the specified pre-pandemic error structure, rather than as a conventional confirmatory p-value. Finally, the approximate nine-month shift used for seasonality is descriptive and ignores gestational variation.

## Implications for future research

Future work using Brazilian SINASC microdata could test whether the July 2021 signal is concentrated among singleton births or maternal and geographic subgroups, and whether fetal survival or pregnancy composition contributes to the national pattern. The later observations also suggest prospectively specified tests: geographic and subgroup coherence after Pelé’s death; conception-cohort analyses around the end of the COVID-19 emergency; and whether June 2024 SRB deviations track dengue intensity or maternal dengue, with sex-specific fetal survival assessed rather than assumed [40]. Exposure definitions and lag windows should be specified in advance wherever possible.

## Conclusions

Brazilian national data show a transient SRB decline in July 2021, three months after the April 2021 Gamma-wave mortality peak. Concordant downward deviations across all five macroregions, largest in the North, add geographic coherence to this prespecified temporal association. The broader series also identified hypothesis-generating deviations: a February 2023 increase nine months after Brazil ended its COVID-19 public-health emergency, compatible with a behavioural pathway involving increased sexual intercourse around conception, and an April 2023 decline approximately four months after Pelé’s death, paralleling a reported 4–5-month decline after the death of Diana, Princess of Wales [37]. These retrospective correspondences do not establish event effects. Together, the findings highlight SRB’s population-level utility and warrant investigation of stress-related fetal survival and behavioural pathways; the ecological design precludes causal inference.

## Data Availability

All data produced are available online at https://tabnet.datasus.gov.br/cgi/deftohtm.exe?sinasc/cnv/nvuf.def

## Statements and Declarations

## Author Contributions

All authors (DFBL, RM, PTC, VG, WM, GM, and GL) conceived the study. DFL identified the relevant publicly available data sources and contributed her expertise on obstetric practice and birth patterns in Brazil. GM conducted the statistical analyses with input from all authors. All authors contributed to the interpretation of the results. GM and GL prepared the first draft of the manuscript. All authors critically reviewed and revised the manuscript and approved the final version.

## Ethics Approval

This study used publicly available, aggregate live-birth data from Brazil’s *Sistema de Informações sobre Nascidos Vivos* (SINASC), without individual-level or identifiable information. Ethics committee approval was not required for this study.

## Consent to Participate

Not applicable. The study used publicly available, aggregate data and did not involve direct participation of human subjects.

## Consent to Publish

Not applicable. The study did not use individual-level or identifiable information.

## Data Availability

The live-birth data analysed in this study are publicly available from Brazil’s *Sistema de Informações sobre Nascidos Vivos* (SINASC), Ministry of Health: https://tabnet.datasus.gov.br/cgi/deftohtm.exe?sinasc/cnv/nvuf.def

